# Hospitalisation for acute heart failure and in-hospital mortality before, during, and after the COVID-19 pandemic in France: A Nationwide cohort study from 2013 to 2024

**DOI:** 10.1101/2025.04.14.25325820

**Authors:** P Moulaire, T Delory, S Oghina, T Damy, M Espagnacq, M Khlat, S Le Cœur, G Hejblum, N Lapidus, the COVID-HOSP study group

## Abstract

**Introduction:** Healthcare systems were reorganised in 2020 to manage the COVID-19 pandemic. Despite their urgent status, hospital admissions for acute heart failure (AHF) were reported to decline from 9% to 66% worldwide between 2020 and 2021, with divergent findings regarding in-hospital mortality. This study aimed to investigate in detail the evolution of AHF hospitalisations and in-hospital mortality in France from 2013 to 2024.

**Methods:** Based on the 2.9 million AHF hospitalisations recorded in France from 2013 to 2024, yearly numbers of hospitalisations and deaths expected in years 2020 to 2024 were estimated using a Poisson regression model, with 2013–2019 as the reference period. The differences between observed and expected event counts in the years 2020 to 2024 were used to quantify the disruptions that occurred since the emergence of the pandemic.

**Results:** A total deficit of −222,913 [−223,908; −221,926] (mean [95% CI]) AHF hospitalisations was estimated for the 2020-2024 years, corresponding to a 16.1% decrease compared to pre-pandemic trends. The yearly reduction in AHF hospitalisations worsened over time, from −39,268 [−39,685; − 38,847] fewer cases in 2020 to −55,521 [−55,984; −55,051] in 2024. In parallel with the decline in AHF hospitalisations, estimated excess in-hospital deaths were 828 [729; 928], 1,625 [1,517; 1,731], 2,427 [2,323; 2,531], 1,739 [1,634; 1,844], and 1,175 [1,068; 1,281] for the years 2020 to 2024, respectively. These correspond to relative increases in in-hospital mortality ranging from 4.4% to 13.2% compared to expected values. The disruptions in both hospital admissions and in-hospital mortality affected more females than males.

**Conclusions:** The apparent long-lasting changes in the management of AHF patients in France observed since the COVID-19 pandemic emergence, particularly among females, suggest improving the preparedness for future crises and require addressing the current sustained disruptions.

**Key Messages:** *What is already known on this topic:* In 2020 and 2021, hospitalisations for acute heart failure were reported to decline worldwide following the onset of the COVID-19 pandemic. However, findings on concomitant in-hospital mortality have remained unclear, and little is known about whether these disruptions persisted through 2022 to 2024.

*What this study adds:* Analyses of exhaustive French national data indicate that the decline in admissions observed in 2020 persisted and even worsened through 2024, with an overall decrease of 16.1%. In parallel, in-hospital mortality was estimated in each year from 2020 to 2024, and the resulting excess corresponded to a cumulative increase of 8.4%. Females were more impacted than males by both disruptions.

*How this study might affect research, practice or policy:* This study highlights critical warnings on ongoing disruptions affecting patients hospitalised for acute heart failure in France and identifies the subpopulations most impacted. These findings might contribute to guide targeted mitigation strategies and to enhance the preparedness of national health systems for future health crises.

## Introduction

At the end of 2019, COVID-19 emerged in Wuhan, China,[1] and by January 30, 2020, the World Health Organization (WHO) declared it a Public Health Emergency of International Concern (PHEIC) due to its rapid global spread.[2] In response to the crisis, many countries implemented unprecedented measures, including lockdowns and significant reorganisation of healthcare systems.[3] In France, non-urgent hospitalisations were deferred to mitigate the virus spread and prioritise the care of COVID-19 patients.[4] However, despite the urgent nature of acute heart failure (AHF), a decline in hospitalisations for this condition was observed early in the pandemic period, in March 2020.[5] A decrease in the incidence of AHF hospitalisations, not only in 2020, but also in 2021 and 2022, was highlighted in a more recent study published in 2024, which focused on the epidemiology of hospitalised heart failure in France.[6] However, this latter study considered only the first hospitalisation of the year without any prior hospitalisations in the preceding five years and did not provide precise quantification of either the decrease in hospitalisations or in-hospital mortality in the pandemic period. Several studies have reported similar reductions in AHF hospital admissions across Europe[7,8] and worldwide,[9–11] with decreases ranging from 9% to 66% between 2020 and 2021. However, the reported in-hospital mortality of AHF admissions during the pandemic period varied widely, with some studies reporting an increased mortality,[12–14] while others reported no significant change.[15–17] Additionally, little is known about the persistence of the disruptions, including trends in hospitalisation rates and mortality in 2023 and 2024, while the WHO declared the end of the phase of PHEIC on May 5, 2023.[18]

Given the uncertainties regarding the extent of the decline in AHF hospitalisations, the corresponding in-hospital mortality rates, and the evolution of these patterns in 2023 and 2024, this study aimed to provide detailed investigations of the disruptions during and after the pandemic period in France. Hospitalisation data from 2013 to 2019 were analysed and used as a reference to model the historical evolution in the numbers of AHF hospitalisations and corresponding in-hospital deaths, allowing for estimates of expected values from 2020 to 2024 in the absence of the pandemic. The primary metrics to quantify the disruptions occurring during the 2020–2024 period were the differences between observed and expected hospitalisations and deaths. One may hypothesise that the decline in AHF admissions observed during the early phase of the pandemic has likely tapered as the pandemic’s intensity decreased, potentially with a catch-up phenomenon. By 2023 and into 2024, a return to pre-pandemic trends might have been expected. Hypothesising what should be the evolving trends in corresponding in-hospital mortality is more challenging. On the one hand, hospital admissions may have been restricted to the most severe emergencies, which could have potentially increased in-hospital mortality. On the other hand, it is possible that more critical patients succumbed to COVID-19, leaving fewer severe AHF cases, which could lower in-hospital mortality. In any scenario, we expect mortality rates to return, at most, to pre-pandemic levels after 2023.

In summary, the overall objective of this study was to provide a comprehensive overview of the evolution in the management of AHF patients over the past 12 years and to evaluate the full impact of the COVID-19 pandemic period on hospitalisations and associated mortality in France, from 2020 to 2024.

## Materials and Methods

### Data sources

Data on hospitalisations for AHF come from the French Hospital Discharge Database (PMSI), which is part of the National Health Data System (SNDS). The SNDS is a national claims database designed to reimburse care via the French Health Insurance System, covering nearly 100% of the population receiving healthcare in France.[19] With the growing interest in the scientific medical community for real-world data derived from large administrative healthcare databases, the SNDS has, for instance, previously been used to study the impact of the pandemic period on acute cardiovascular diseases[20–22] and other diseases.[23,24] Data on the French population structure were also used in this study and are available in open access from the French National Institute for Statistics and Economic Studies (Insee).[25] There was no missing variable value in our database queries, and therefore, analyses assumed the absence of missing data.

### Population under study

This open cohort study, covering the entire French population from 1 January 2013 to 31 December 2024, is reported following the STROBE guidelines[26]. As the age structure of the French population is reported on January 1 of each year, the age considered for participants was that on January 1, and data were aggregated by year. Patients aged 99 years or more were handled in a single category. Following the SAGER guidelines[27], sex was always taken into account in the study design and report.

### Outcomes of interest

Two outcomes were identified and analysed during the study period: the annual number of AHF hospitalisations and the number of associated in-hospital deaths. AHF hospitalisation events were identified in the SNDS database based on the International Classification of Diseases, 10^th^ Revision (ICD-10), using an algorithm developed by the Healthcare Expenditures and Conditions Mapping[28] (see Supplemental Text 1). In-hospital death was defined as an AHF hospital stay that ended with the death of the patient. Deaths attributed to COVID-19 among patients who died during hospitalisation for AHF were assessed in a supplemental analysis (see Supplemental Analysis 1).

### Analysing the evolution of the numbers of AHF hospitalisations and associated in-hospital deaths

To quantify the impact of the pandemic period on the outcomes of interest, a two-step approach was used, comparing the expected and observed numbers of AHF hospitalisations and associated in-hospital deaths. In the first step, the pre-pandemic period (2013 to 2019) was used as a reference to model the natural evolution of both outcomes just before the pandemic onset. The modelling was based on a Poisson regression, detailed in the next subsection. In the second step, for each outcome of interest, the corresponding regression model was used to predict the expected values for years 2020 to 2024, assuming the natural evolution observed during the pre-pandemic period had not been disrupted by the pandemic. The expected values were then compared to the observed occurrences, enabling us to characterise how the natural evolution of AHF hospitalisations and corresponding in-hospital deaths were disrupted following the emergence of the COVID-19 pandemic (see Supplemental Figure 1). Similar models have been previously applied to estimate expected mortality levels for the calculation of cause-specific excess mortality[29] or all-cause excess mortality.[30,31]

### Statistical analysis

A Poisson regression model was used to estimate the numbers of expected AHF hospitalisations and in-hospital deaths after the arrival of the pandemic, considering pre-pandemic years 2013−2019 as the reference period.

The model included age as a year-specific categorical variable to accurately capture potential non-linear effects of age on the numbers of hospitalisations and in-hospital deaths, an interaction between age and sex to precisely estimate sex differences across age groups, and calendar year as a numeric variable to assess yearly temporal trends in the numbers of hospitalisations and deaths. To estimate expected AHF hospitalisations while accounting for changes in population size and age structure over the study period, the natural logarithm of the population was used as an offset term. For in-hospital mortality among AHF admissions, the number of AHF admissions was used as an offset term to account for changes in hospital load over time. The Akaike Information Criterion (AIC) was employed as a selection criterion to optimise goodness-of-fit and avoid overfitting (Supplemental Table 2). To investigate seasonal periodicity, additional analyses were performed at both monthly and weekly temporal resolutions, employing sinusoidal and spline-based models. The details of these models, along with graphical visualisations of the predicted trends and their impact on the final estimates, are available in the Supplemental Material (Supplemental Analysis 2, Supplemental Figures 2 to 4). The Poisson regression used for modelling the number of hospitalisations was:

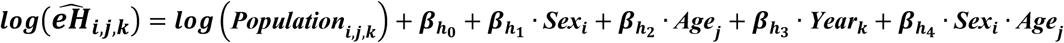

With *H_i_*_,*j*,*k*_ the number of observed AHF hospitalisations (in the SNDS database) during year *k* (*k ε* {2020, 2021, 2022, 2023,2024}) in the stratum of persons with sex *i* and age *j* and 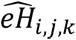 the corresponding number of AHF hospitalisations expected to occur with the model during year *k* in the same population stratum.

The resulting difference between observed and expected values is 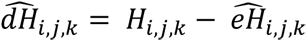, and the disruption observed year k compared to the expected natural evolution of the number of AHF hospitalisations was calculated as follows:

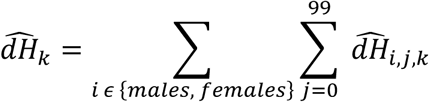

Similarly, the Poisson regression used for modelling the number of in-hospital deaths among AHF hospitalisations was:

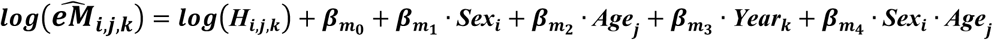

and the disruption observed year *k* compared to the expected natural evolution of the number of in-hospital deaths was calculated as follows:

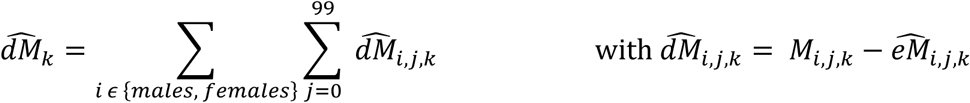

All analyses were performed with statistical software R 4.0.2 (R Foundation for Statistical Computing, Vienna, Austria), package ggplot2 was used to generate figures, package stats was used for the generalised linear models, and 95% confidence intervals (CIs) were estimated over 10,000 bootstrap replications.

## Results

### Population under study

Between 2013 and 2024, the French population increased from 65.6 million to 68.4 million, and the median age rose from 40 to 42 years (see study flow diagram in Figure 1). During the study period, which included a total of 805,627,212 person-years, 2,922,319 AHF hospitalisations occurred at 83 [74; 88] years old (median age [interquartile range]), including 246,080 corresponding in-hospital deaths at 86 [80; 91] years old.

**Figure 1:**
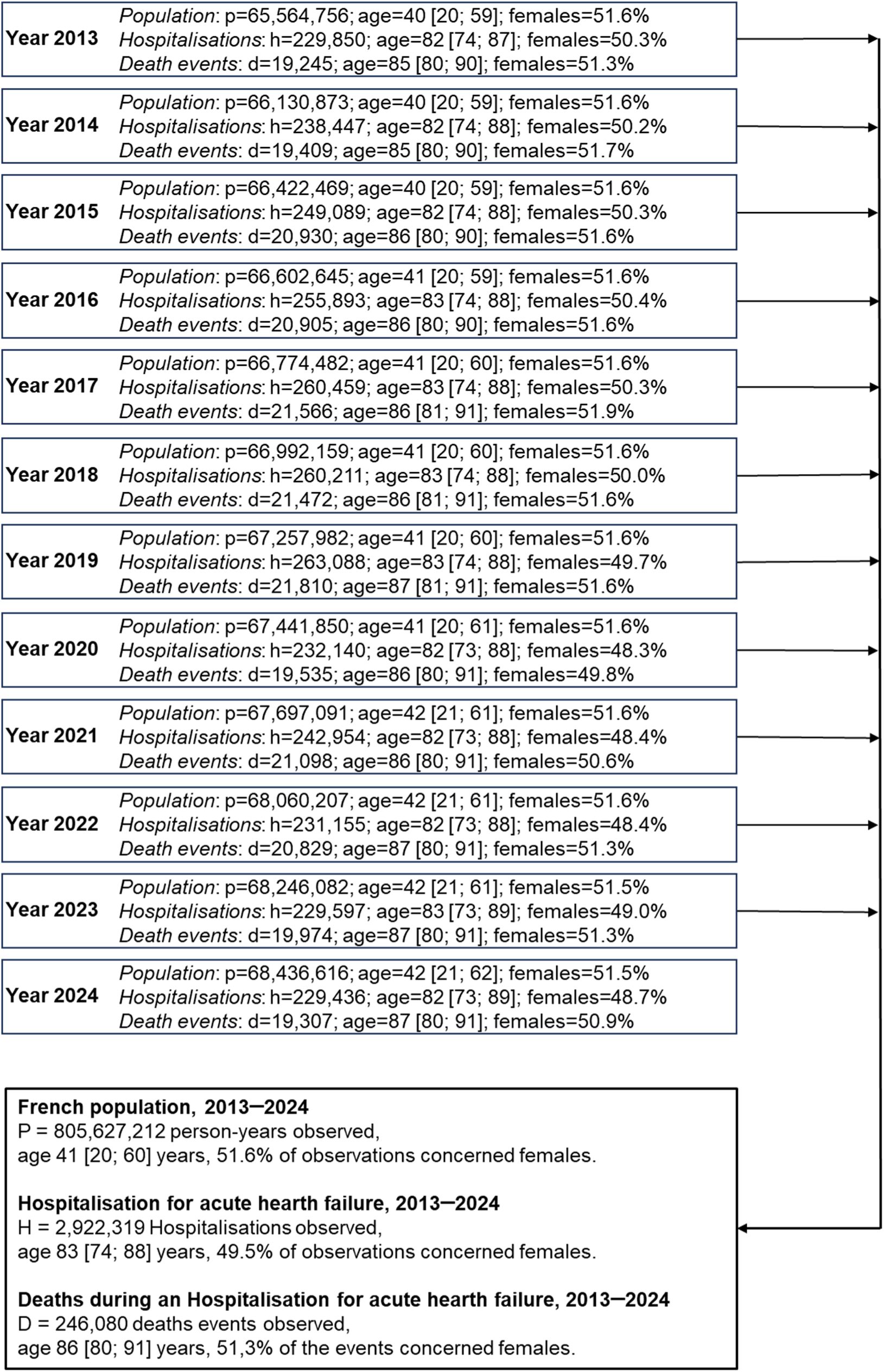
study flow diagram. Abbreviations used: p corresponds to the population size on January 1 of this given year, h corresponds to the total number of AHF hospitalisations and d to the total number of death events that occurred in France each year. Age is reported by its median value [interquartile range], and the percentage of data concerning females is also reported.

Figure 2 presents the number of AHF hospitalisations that occurred each week over the study period, with the corresponding distribution by age and sex. This synthesis picture of exhaustive National data in France over time provides key insights into AHF hospitalisation trends. Firstly, hospitalisations predominantly involved patients aged between 70 and 95 years. Secondly, data present seasonal patterns, with a peak at the beginning of each year and a decrease during summer months. Thirdly, the number of AHF hospitalisations gradually increased from 2013 to 2019. Finally, a sharp decline in hospital admissions was notable during the first wave of the pandemic in April 2020, followed by a series of fluctuations through to 2024, without ever returning to pre-pandemic levels. These visual trends are corroborated by the numerical data in the flow diagram (see Figure 1).

**Figure 2.**
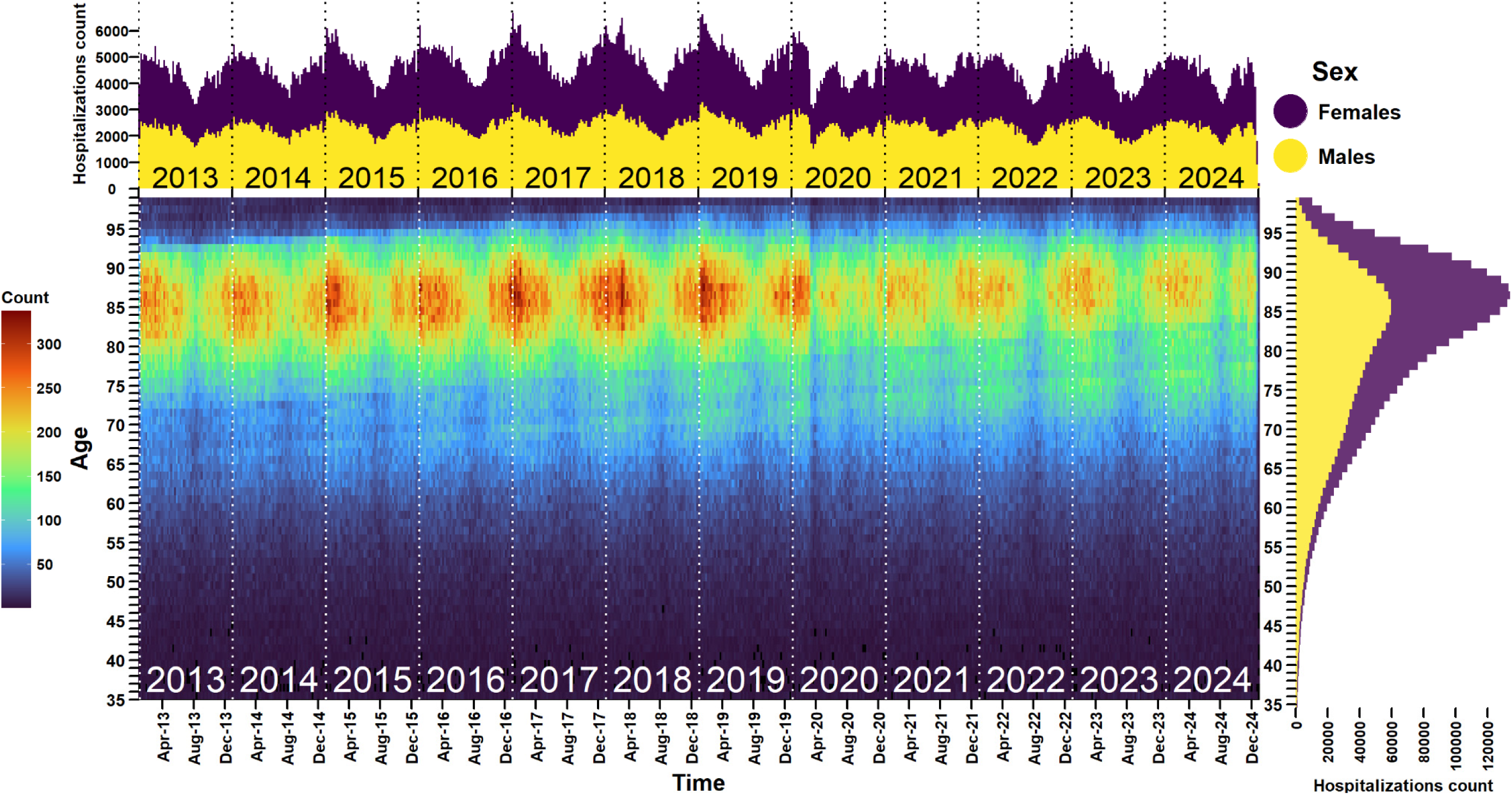
Occurrences of AHF hospitalisations observed each week in males and females according to age in France, years 2013 to 2024 (N = 2,922,319) Main panel: Weekly number of AHF hospitalisations according to age. For example, in January 2019, the colour gradient is red for individuals aged between 85 and 90, corresponding to a weekly number of around 300 hospitalisations. Top panel: Cumulative numbers (in males and females) of AHF hospitalisations each week in France from 2013 to 2024, broken down by sex. Right panel: Cumulative number (over time) of AHF hospitalisations according to age in males and females.

### Evolution in the number of AHF hospitalisations

As illustrated in Figure 3, the numbers of AHF hospitalisations observed in years 2020 to 2024, i.e., after the emergence of the pandemic, consistently remained below the expected numbers based on the trend from the previous seven pre-pandemic years (2013–2019), for both males and females. The decline pattern intensified over time, starting with −39,268 [−39,685; −38,847] fewer hospitalisations than expected in 2020, and peaking at −55,521 [−55,984; −55,051] fewer in 2024. These estimates correspond to annual reductions ranging from −11.3% to −19.5%. Considering the overall pandemic period, there were −222,913 [−223,908; −221,926] fewer AHF hospital admissions, representing a −16.1% reduction from the expected figures. Females were more affected than males each year, with a total of −117,459 [−118,142; −116,774] fewer hospitalisations than expected (−17.2%) compared to −105,454 [−106,169; −104,755] fewer hospitalisations (−15.0%) for males between 2020 and 2024. Detailed annual estimates of the decline in AHF hospitalisations, stratified by sex, are provided in Table 1. As shown in Supplemental Figure 5, the drop in hospitalisations involved patients aged 83 [77; 88] years between 2020 and 2024, while the median age of hospitalised patients during the same period was 82 [74; 88] years.

**Figure 3.**
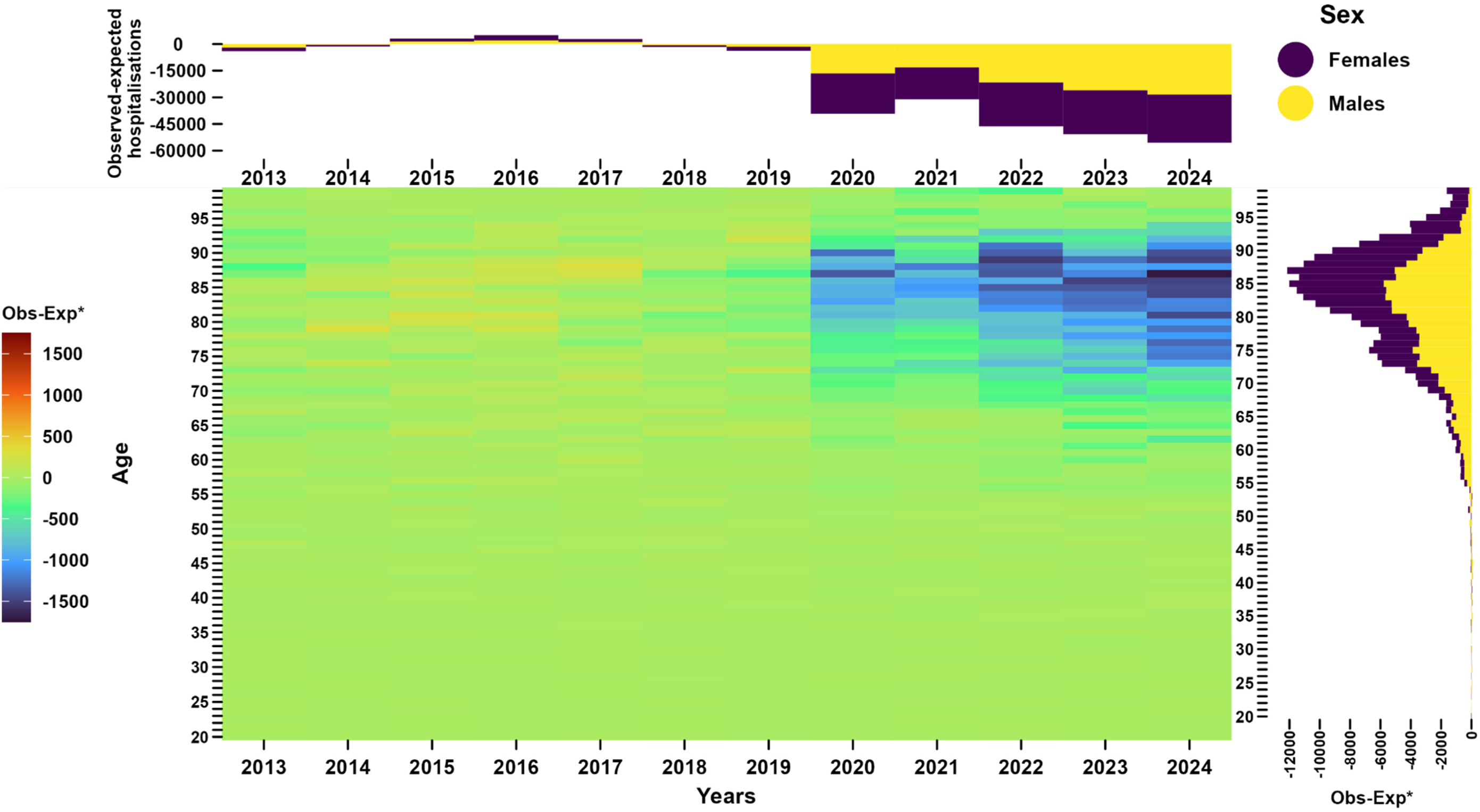
Differences between observed and expected numbers of AHF hospitalisations, France, 2013–2024. *Observed minus expected Main panel, annual difference between observed and expected hospitalisations according to age. For example, in 2024, the colour gradient is dark blue for individuals aged 85, corresponding to 1,500 fewer admissions than expected. Top panel, annual cumulative (in males and females) difference between observed and expected hospitalisations, years 2013 to 2019 were used as the reference period. Right panel, cumulative (over time, years 2020 to 2024) difference between observed and expected hospitalisations according to age and sex.

**Table 1:**
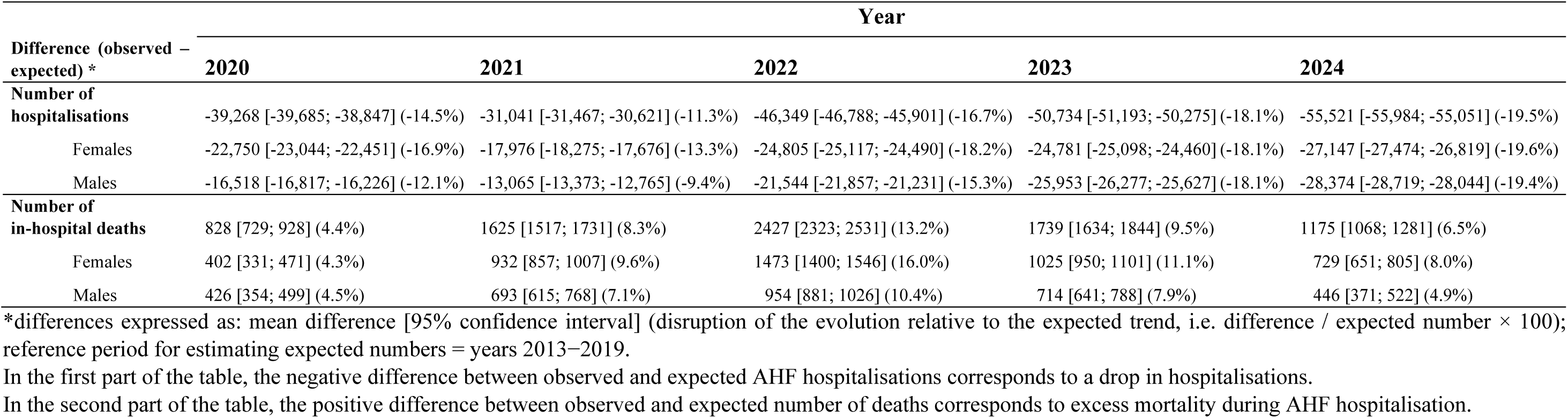
Differences (observed - expected) in the numbers of hospitalisations for acute heart failure, and in the numbers of corresponding in-hospital deaths, France, 2020−2024.

### Evolution in the number of in-hospital deaths among AHF hospitalisations

Figure 4 illustrates the evolution of in-hospital mortality among patients hospitalised for AHF in the years 2020 to 2024. Throughout the overall pandemic period, in-hospital mortality exceeded expectations for both males and females. This excess mortality steadily increased from 828 [729; 928] excess deaths in 2020 to 2,427 [2,323; 2,531] in 2022, before decreasing to 1,175 [1,068; 1,281] in 2024. These excess deaths represented relative increases ranging from 4.4% to 13.2% compared to the pre-pandemic trend. The cumulative toll over the years 2020 to 2024 reached 7,794 [7,557; 8,028] excess deaths (8.4%). Since 2021, the excess in-hospital mortality has affected more females than males, with corresponding relative increases ranging from 9.6% to 16.0% for females, compared to 7.1% to 11.1% for males. Detailed annual estimates by sex are provided in Table 1. As shown in Supplemental Figure 6, patients contributing to the excess in-hospital mortality had a median age of 86 years [78; 91], compared to 87 years [80; 91] for those observed.

**Figure 4.**
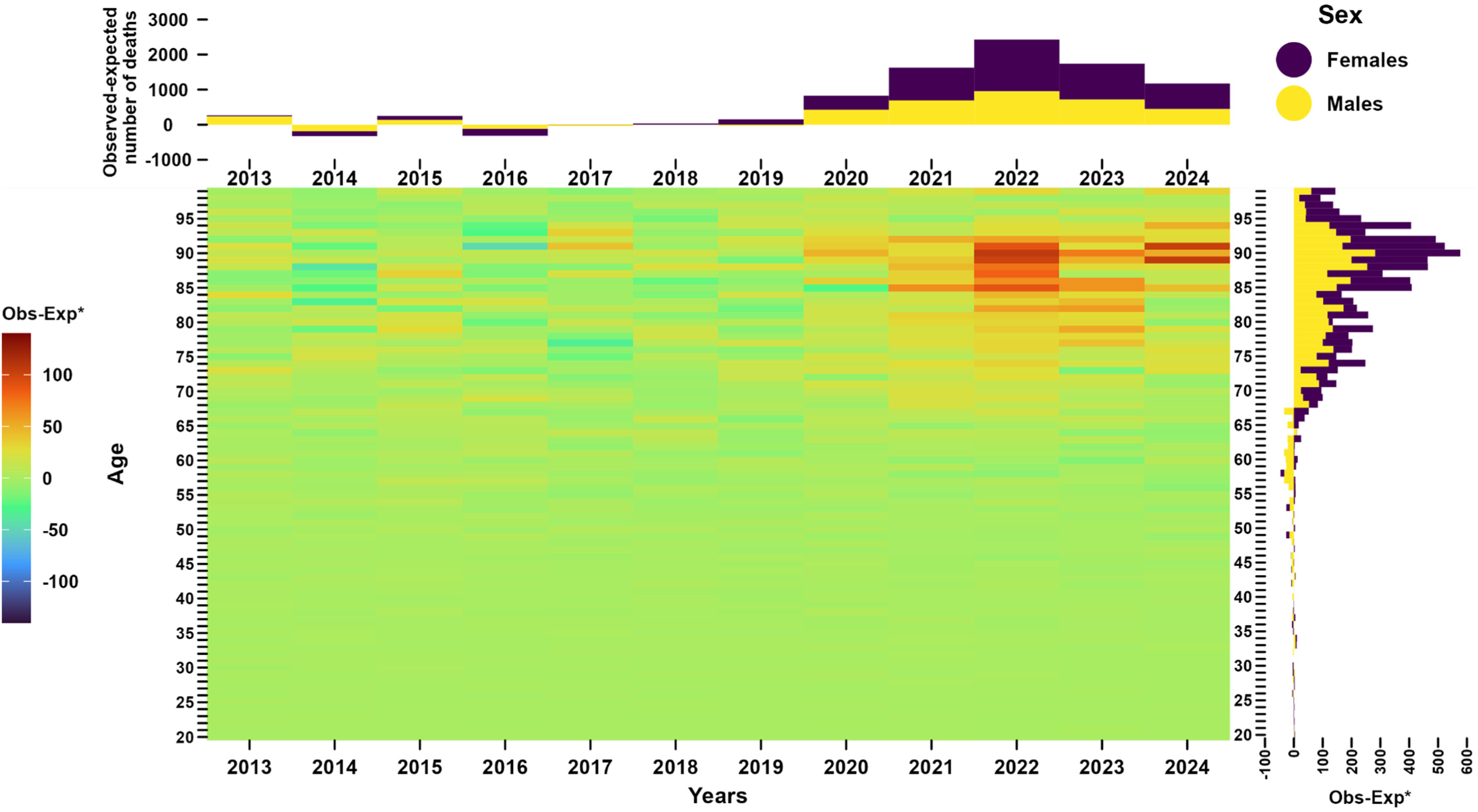
Differences between observed and expected numbers of in-hospital deaths, France, 2013–2024. *Observed minus expected Main panel, annual difference between observed and expected in-hospital deaths according to age. For example, in 2022, the colour gradient is red for individuals aged 85, corresponding to 100 excess in-hospital deaths. Top panel, annual cumulative (in males and females) difference between observed and expected in-hospital deaths, years 2013 to 2019 were used as the reference period. Right panel, cumulative (over time, years 2020 to 2024) difference between observed and expected in-hospital deaths

## Discussion

### Key results

In this open cohort study of the French population from 2013 to 2024 (805,627,212 person-years), trends in AHF hospitalisations and corresponding in-hospital mortality were analysed to accurately quantify the extent of disruption during and after the COVID-19 pandemic period (2020–2024). The study provides evidence of a major and prolonged disruption since the onset of COVID-19, which remained substantial in 2024, even after the end of the pandemic. Specifically, a 14.5% decline in hospital admissions occurred in 2020, and, surprisingly, this decline worsened to 19.5% in 2024. The decrease, observed throughout the pandemic period and persisting even in 2024, affected both sexes but was more pronounced among females. As shown in Supplemental Figure 5, the decline in hospitalisations was more pronounced among older patients. This trend is further supported by the flow diagram, which illustrates that the median age of hospitalised patients increased from 2013 to 2019, reflecting the ageing population in France. However, during the pandemic period, a decrease in the median age of patients hospitalised for AHF was observed, indicating that the decline in hospitalisations particularly affected the oldest patients. As shown in Supplemental Analysis 3, the decline in hospitalisations appears to be specific to AHF. Indeed, while the healthcare system experienced global disruptions since 2020, the reduction in hospitalisations was even more pronounced in AHF patients compared to the general population, and this gap widened over time (Supplemental Figure 7, Supplemental Table 3).

In contrast to the decline in AHF hospitalisations, in-hospital mortality exceeded expectations, with excess mortality ranging from 4.4% to 13.2% between 2020 and 2024. This corresponded to an estimated total of 7,794 [7,557; 8,028] excess in-hospital deaths, representing an 8.4% increase compared to the expected mortality based on the pre-pandemic trend. Similar to the decline in hospital admissions, the increase in in-hospital mortality affected both sexes throughout the pandemic, and after, and was more pronounced in females. As illustrated in Supplemental Figure 6, the rise in in-hospital mortality extended beyond the oldest patient groups.

### Interpretation

The study confirms previous national observations regarding the decline in AHF hospitalisations between 2020 and 2022, while providing more detailed insights into the magnitude of the disruption. Additionally, it extends the analysis to include 2023 and 2024, offering a comprehensive assessment of the pandemic period’s long-term impact thanks to nationwide data spanning 12 consecutive years. The drop in hospitalisations, ranging from 11.3% to 14.5% in 2020 and 2021, aligns with figures reported in Europe[7,8] and worldwide.[9–11] Importantly, while it was anticipated that after the widespread vaccination against COVID-19, and the reduction in pandemic severity from 2022 to 2024, the situation would return to the pre-pandemic state, the study evidenced that the disruption worsened in the following years, peaking at 19.5% fewer hospitalisations in 2024.

Two contrasting effects of the pandemic may have contributed to the substantial decline in AHF hospitalisations. Firstly, a series of protective effects related to the pandemic period could have led to a reduction in AHF incidence and, consequently, fewer hospitalisations. Key changes in population behaviours since 2020, such as widespread mask usage, have reduced the transmission of respiratory viruses, which are well-known to increase the risk of acute decompensation in AHF patients.[32,33] Shifts in healthcare practices, including the rapid expansion of telemedicine,[34,35] the introduction of new medications,[36] and major initiatives by the National Health Insurance since 2022,[37] may have improved patient care and further contributed to the reduction in hospitalisations for AHF. While these factors likely explain part of the decrease in hospitalisations, they do not fully account for the abrupt and persistent nature of this change, nor do they clarify why older individuals and females were particularly affected.

Secondly, a series of detrimental effects from the pandemic may have led to premature deaths before hospitalisation, thus reducing the overall number of hospitalisations. The widespread impact of the pandemic period resulted in a sudden and prolonged period of excess mortality in France,[38,39] which is consistent with the observed decline in hospital admissions for the years 2020 to 2022. The major risk factors for COVID-19 mortality, such as older age and comorbidities,[40] closely match the profile of patients typically hospitalised for AHF. However, this direct effect of the pandemic cannot fully explain the decrease in AHF hospitalisations. COVID-19-related deaths officially decreased from 69,000 in 2020[41] to 5,572 in 2023,[42] while the present study results indicate that the decline in the number of AHF hospitalisations was 39,268 fewer admissions in 2020, and increased to 50,734 in 2023. More importantly, in the post-pandemic period, in 2024, although the number of officially reported COVID-19-related deaths dropped to just 96,[43] the decline in AHF hospitalisations reached its peak, with 55,521 fewer admissions. Beyond direct competitive effects, some negative indirect consequences of the pandemic may also have contributed to premature death.[44] Factors such as fear of seeking hospital care, or reduction of the initiation of medication treatments for cardiovascular risk documented in France in 2020[45] may have led to out-of-hospital AHF-related deaths before hospitalisation. Indeed, studies have reported a significant increase in out-of-hospital AHF-related deaths occurring at home during 2020, with rises ranging from 25% to 35% compared to the pre-pandemic period.[46,47] Numerous studies have documented gender inequality in the management of cardiovascular care,[48–50] making it likely that, during a crisis, females were among the first to suffer the negative consequences. Notably, a rise in cardiovascular mortality was observed in France from 2020 to 2022, particularly among females.[51]

Study results indicate that in-hospital deaths among AHF hospitalisations exceeded expected levels throughout the pandemic. While some previous studies found no significant change in in-hospital mortality,[12–14] others reported an increase.[15–17] These discrepancies may be due to differences between countries or to the greater statistical power provided by the exhaustive data used in the present study. The increase in in-hospital mortality during the early phase of the pandemic could be attributed to COVID-19-related deaths. In 2020, for instance, 929 AHF-hospitalised patients died of COVID-19, potentially explaining the 828 excess deaths estimated for that year (Supplemental Table 1). However, COVID-19 mortality among AHF patients predominantly affected males and has declined over time. In contrast, in-hospital excess mortality has risen, particularly among females. Notably, in 2022, COVID-19 mortality could account for only 27% of the excess in in-hospital mortality observed in females. While COVID-19-related mortality, as a new cause of death emerging in 2020, could have explained the increase in in-hospital mortality among AHF patients, this is not consistent with the worsening mortality over time, nor with the disproportionate impact on females. A reduction in hospital capacity in France,[52] which has worsened since the start of the pandemic, may have restricted hospitalisations to the most severe cases, thereby increasing in-hospital mortality rates. The persistent excess mortality observed in this study is concerning and suggests a profound and long-lasting disruption in the healthcare system’s ability to manage AHF patients, particularly females.

### Strengths and limitations

This study is based on exhaustive data of AHF hospitalisations in France, and it covers not only the entire pandemic period but also one full post-pandemic year. The evolving patterns reported are reliably representative of the whole country. Moreover, the analyses include seven years of pre-pandemic data, allowing historical trends before the onset of the pandemic to be taken into account and enabling reliable estimates of the subsequent perturbations. Most studies published to date have shorter reference periods and/or only estimate disruptions for parts of the pandemic. This study not only offers detailed results on the decline in hospital admissions and the increase in in-hospital mortality by year, sex, and age, but also documents the evolution of these indicators over 12 consecutive years at the national level.

This study has several limitations. The main limitation lies in the lack of access to key clinical and behavioural variables. Important confounding factors such as weight, tobacco use, and blood pressure, as well as clinical information such as biological and radiological results, were not available for analysis. This is due to the nature of the database, which is primarily administrative and does not yet incorporate clinical data. In addition, patient identification relied on ICD-10 codes, which are susceptible to coding errors; however, this method has been previously assessed and validated for the identification of acute heart failure cases[53,54]. Additionally, as healthcare systems vary between countries, the findings of this national study cannot be directly generalised to other nations. However, as previously mentioned, similar patterns have been reported in studies conducted elsewhere. Several hypotheses were proposed regarding the impact of the COVID-19 pandemic on AHF hospitalisations and mortality. These include both protective and negative effects, direct impacts on mortality, and indirect disruptions to the healthcare system. However, due to the complexity and multifactorial nature of these causes, it is not possible to draw causal conclusions. Nevertheless, despite the potential limitations of the study, the large sample size and the magnitude of the estimated disruptions strongly support the conclusion that a major disturbance in the management of AHF patients occurred at the onset of the pandemic and has continued through to 2024.

## Conclusion

This study reveals significant disruptions in the healthcare pathway for AHF, marked by a decline in hospitalisations and an increase in in-hospital mortality from the onset of the COVID-19 pandemic in 2020 through to 2024, with a particular impact on females. Further research devoted to exploring the underlying causes of these persistent disruptions is of primary importance for developing strategies to mitigate their impact.

## Acknowledgements

The COVID-HOSP study group: Tristan Delory (Centre Hospitalier Annecy Genevois, Annecy, France); Fanny Duchaine (IRDES, Paris, France); Maude Espagnacq (IRDES, Paris, France); Gilles Hejblum (INSERM, Paris, France); Myriam Khlat (INED, Aubervilliers, France); Nathanaël Lapidus (INSERM, Paris, France); Sophie Le Cœur (INED, Aubervilliers, France); Elhadji Leye (INSERM, Paris, France); Paul Moulaire (INSERM, Paris FRANCE); Jonas Poucineau (INED, Aubervilliers, France).

## Funding statement

This work was supported by the Initiative Économie de la Santé of Sorbonne Université (Idex Sorbonne Université, programmes Investissements d’Avenir), and by the Ministère de la Solidarité et de la Santé (Programme de Recherche sur la Performance du Système des Soins, PREPS 20-0163). The sponsor and the funders had no role in study design, data collection and analysis, interpretation of data, decision to publish, or preparation of the manuscript.

## Competing Interests statement

The authors of this manuscript have no conflict of interest to disclose.

## Data availability statement

According to the principles of data protection and French regulations, the authors cannot publicly release the data from the French National Health Data System (SNDS). However, any person or structure, public or private, for-profit or non-profit, can access SNDS data upon authorisation from the French Data Protection Office (CNIL, Commission Nationale de l’Informatique et des Libertés) to carry out a study, research, or an evaluation of public interest (https://www.snds.gouv.fr/SNDS/Contexte-et-perspectives-reglementaires#).

## Ethics approval

The SNDS is a set of strictly anonymous databases comprising all mandatory national health insurance reimbursement data. Since June 30, 2021, INSERM has direct access to the SNDS. This permanent access is provided according French Decree No. 2016-1871 of December 26, 2016 relating to the processing of personal SNDS data and French law articles Art. R. 1461-1325 and 14. This study was declared prior to its initiation to the CepiDc-INSERM registry of studies requiring the use of the SNDS. In accordance with national legislation and EU General Data Protection Regulation, written informed consent for participation was not required for this study.

## Author Contributions

TD, GH, and NL initiated the study. GH and NL supervised the study; GH, NL, and PM designed the experimental plan; PM managed data and performed the analyses; GH, NL, and PM can take responsibility for the integrity of the data and the accuracy of the data analysis, PM is the guarantor; GH, NL, and PM prepared the first draft of the manuscript; All authors (TD, SO, TD, ME, MK, SLC, GH, NL and PM) contributed to interpretation of the data, critically revised the manuscript, and approved the final version.

## Patient and public involvement

Patients and/or the public were not involved in the design, conduct, reporting, or dissemination plans of this research.

## Notes

### Competing Interest Statement

The authors have declared no competing interest.

### Funding Statement

This work was supported by the Initiative Economie de la Sante of Sorbonne Universite (Idex Sorbonne Universite, programmes Investissements d'Avenir), and by the Ministere de la Solidarite et de la Sante (Programme de Recherche sur la Performance du Systeme des Soins, PREPS 20-0163). The sponsor and the funders had no role in study design, data collection and analysis, interpretation of data, decision to publish, or preparation of the manuscript.

### Author Declarations

The SNDS is a set of strictly anonymous databases comprising all mandatory national health insurance reimbursement data. Since June 30, 2021 INSERM has direct access to the SNDS. This permanent access is provided according French Decree No. 2016-1871 of December 26, 2016 relating to the processing of personal SNDS data and French law articles Art. R. 1461-1325 and 14. This study was declared prior to its initiation to the CepiDc-INSERM registry of studies requiring the use of the SNDS. In accordance with national legislation and EU General Data Protection Regulation, written informed consent for participation was not required for this study.

### Summary of Updates

Year 2024 has been included in the analysis.

## References

1. Maxmen A. Wuhan market was epicentre of pandemic’s start, studies suggest. Nature. 2022 Feb 27;603(7899):15–6.

2. Statement on the second meeting of the International Health Regulations (2005) Emergency Committee regarding the outbreak of novel coronavirus (2019-nCoV) [Internet]. [cited 2024 Sep 11]. Available from: https://www.who.int/news/item/30-01-2020-statement-on-the-second-meeting-of-the-international-health-regulations-(2005)-emergency-committee-regarding-the-outbreak-of-novel-coronavirus-(2019-ncov)

3. Hsiang S, Allen D, Annan-Phan S, et al. The effect of large-scale anti-contagion policies on the COVID-19 pandemic. Nature. 2020 Aug;584(7820):262–7.

4. Tuppin P, Lesuffleur T, Constantinou P, et al. Underuse of primary healthcare in France during the COVID-19 epidemic in 2020 according to individual characteristics: a national observational study. BMC Prim Care. 2022 Aug 9;23(1):200.

5. Huet F, Prieur C, Schurtz G, et al. One train may hide another: Acute cardiovascular diseases could be neglected because of the COVID-19 pandemic. Arch Cardiovasc Dis. 2020 May 1;113(5):303–7.

6. Olié V, Isnard R, Pousset F, et al. Epidemiology of hospitalized heart failure in France based on national data over 10 years, 2012–2022. ESC Heart Fail [Internet]. [cited 2024 Dec 20];n/a(n/a). Available from: https://onlinelibrary.wiley.com/doi/abs/10.1002/ehf2.15137

7. Khan Y, Verhaeghe N, Devleesschauwer B, et al. The impact of the COVID-19 pandemic on delayed care of cardiovascular diseases in Europe: a systematic review. Eur Heart J - Qual Care Clin Outcomes. 2023 Nov 2;9(7):647–61.

8. Cannata A, Watson SA, Daniel A, et al. Impact of the COVID-19 pandemic on in-hospital mortality in cardiovascular disease: a meta-analysis. Eur J Prev Cardiol. 2021 Jul 23;29(8):1266–74.

9. Fox DK, Waken RJ, Johnson DY, et al. Impact of the COVID-19 Pandemic on Patients Without COVID-19 With Acute Myocardial Infarction and Heart Failure. J Am Heart Assoc. 2022 Mar 15;11(6):e022625.

10. Jayagopa PB, Ramakrishnan S, Mohanan PP, et al. Impact of COVID-19 on heart failure hospitalization and outcome in India - A cardiological society of India study (CSI-HF in COVID 19 times study - “The COVID C-HF study”). INDIAN HEART J. 2023 Oct;75(5):370–5.

11. Seidu S, Kunutsor SK, Cos X, et al. Indirect impact of the COVID-19 pandemic on hospitalisations for cardiometabolic conditions and their management: A systematic review. Prim Care Diabetes. 2021 Aug;15(4):653–81.

12. Cannatà A, Bromage DI, Rind IA, et al. Temporal trends in decompensated heart failure and outcomes during COVID-19: a multisite report from heart failure referral centres in London. Eur J Heart Fail. 2020 Dec;22(12):2219–24.

13. König S, Hohenstein S, Meier-Hellmann A, et al. In-hospital care in acute heart failure during the COVID-19 pandemic: insights from the German-wide Helios hospital network. Eur J Heart Fail. 2020 Dec;22(12):2190–201.

14. Kubica J, Ostrowska M, Stolarek W, et al. Impact of COVID-19 pandemic on acute heart failure admissions and mortality: a multicentre study (COV-HF-SIRIO 6 study). ESC Heart Fail. 2022 Feb;9(1):721–8.

15. Wang SY, Seghieri C, Vainieri M, et al. Changes in Acute Myocardial Infarction, Stroke, and Heart Failure Hospitalizations During COVID-19 Pandemic in Tuscany-An Interrupted Time Series Study. Int J Public Health. 2022;67:1604319.

16. Andersson C, Gerds T, Fosbøl E, et al. Incidence of New-Onset and Worsening Heart Failure Before and After the COVID-19 Epidemic Lockdown in Denmark: A Nationwide Cohort Study. Circ Heart Fail. 2020 Jun;13(6):e007274.

17. Cedrone F, Di Martino G, Di Giovanni P, et al. Reduction in Hospital Admissions for Cardiovascular Diseases (CVDs) during the Coronavirus Disease 2019 (COVID-19) Pandemic: A Retrospective Study from a Southern Italian Region in the Year 2020. Healthcare. 2022 May 9;10(5):871.

18. World Health Organization. Statement on the fifteenth meeting of the IHR (2005) Emergency Committee on the COVID-19 pandemic [Internet]. [cited 2024 Jun 28]. Available from: https://www.who.int/news/item/05-05-2023-statement-on-the-fifteenth-meeting-of-the-international-health-regulations-(2005)-emergency-committee-regarding-the-coronavirus-disease-(covid-19)-pandemic

19. Tuppin P, Rudant J, Constantinou P, et al. Value of a national administrative database to guide public decisions: From the systeme national d’information interregimes de l’Assurance Maladie (SNIIRAM) to the systeme national des donnees de sante (SNDS) in France. Rev Epidemiol Sante Publique. 2017 Oct;65:S149–67.

20. Grave C, Gabet A, Puymirat E, et al. Myocardial infarction throughout 1 year of the COVID-19 pandemic: French nationwide study of hospitalization rates, prognosis and 90-day mortality rates. Arch Cardiovasc Dis. 2021 Dec;114(12):768–80.

21. Gabet A, Grave C, Tuppin P, et al. Changes in the epidemiology of patients hospitalized in France with deep venous thrombosis and pulmonary embolism during the COVID-19 pandemic. Thromb Res. 2021 Nov 1;207:67–74.

22. Gabet A, Grave C, Tuppin P, et al. Impact of the COVID-19 pandemic and a national lockdown on hospitalizations for stroke and related 30-day mortality in France: A nationwide observational study. Eur J Neurol. 2021;28(10):3279–88.

23. Leye E, Delory T, El Karoui K, et al. Direct and indirect impact of the COVID-19 pandemic on the survival of kidney transplant recipients: A national observational study in France. Am J Transplant. 2024 Mar 1;24(3):479–90.

24. Poucineau J, Khlat M, Lapidus N, et al. Impact of the COVID-19 Pandemic on COPD Patient Mortality: A Nationwide Study in France. Int J Public Health. 2024 Feb 1;69:1606617.

25. Institut national de la statistique et des études économiques (French National Institute of Statistics and Economic Studies). Séries Population et structure de la population | Insee [Internet]. [cited 2024 Sep 18]. Available from: https://www.insee.fr/fr/statistiques/series/103088458

26. Vandenbroucke JP, Elm E von, Altman DG, et al. Strengthening the Reporting of Observational Studies in Epidemiology (STROBE): Explanation and Elaboration. PLOS Med. 2007 Oct 16;4(10):e297.

27. Heidari S, Babor TF, De Castro P, et al. Sex and Gender Equity in Research: rationale for the SAGER guidelines and recommended use. Res Integr Peer Rev. 2016 May 3;1(1):2.

28. Méthode [Internet]. [cited 2024 Sep 20]. Available from: https://www.assurance-maladie.ameli.fr/etudes-et-donnees/par-theme/pathologies/cartographie-assurance-maladie/methode-cartographie-pathologies-depenses-assurance-maladie

29. Alicandro G, La Vecchia C, Islam N, et al. A comprehensive analysis of all-cause and cause-specific excess deaths in 30 countries during 2020. Eur J Epidemiol. 2023 Nov 1;38(11):1153–64.

30. Barnard S, Chiavenna C, Fox S, et al. Methods for modelling excess mortality across England during the COVID-19 pandemic. Stat Methods Med Res. 2022 Sep;31(9):1790– 802.

31. Fantin R, Barboza-Solís C, Hildesheim A, et al. Excess mortality from COVID 19 in Costa Rica: a registry based study using Poisson regression. Lancet Reg Health – Am [Internet]. 2023 Apr 1 [cited 2024 Jun 28];20. Available from: https://www.thelancet.com/journals/lanam/article/PIIS2667-193X(23)00025-X/fulltext

32. Girerd N, Chapet N, Roubille C, et al. Vaccination for Respiratory Infections in Patients with Heart Failure. J Clin Med. 2021 Jan;10(19):4311.

33. Alon D, Stein GY, Korenfeld R, et al. Predictors and Outcomes of Infection-Related Hospital Admissions of Heart Failure Patients. PLOS ONE. 2013 Aug 23;8(8):e72476.

34. Berthelot E, Flécher E, Roubille F, et al. Impact of the COVID-19 pandemic on the burden of chronic heart failure patients in France. Ann Cardiol Angéiologie. 2021 Oct 1;70(4):191–5.

35. Severino P, Prosperi S, D’Amato A, et al. Telemedicine: an Effective and Low-Cost Lesson From the COVID-19 Pandemic for the Management of Heart Failure Patients. Curr Heart Fail Rep. 2023 Oct 1;20(5):382–9.

36. BrJCardiol. Drug therapy in heart failure – an update from the 2021 ESC heart failure guideline [Internet]. [cited 2024 Sep 19]. Available from: https://bjcardio.co.uk/2022/07/drug-therapy-in-heart-failure-an-update-from-the-2021-esc-heart-failure-guideline/

37. Campagne insuffisance cardiaque [Internet]. [cited 2024 Sep 17]. Available from: https://www.assurance-maladie.ameli.fr/qui-sommes-nous/action/campagnes-communication/campagne-insuffisancecardiaque

38. Blanpain N. 53,800 more deaths than expected in 2022: higher excess mortality than in 2020 and 2021. Insee Première. 2023; 1951 [Internet]. [cited 2024 Sep 19]. Available from: https://www.insee.fr/en/statistiques/7635866

39. Moulaire P, Hejblum G, Lapidus N. Excess mortality and years of life lost from 2020 to 2023 in France: a cohort study of the overall impact of the COVID-19 pandemic on mortality. BMJ Public Health [Internet]. 2025 Mar 4 [cited 2025 Mar 13];3(1). Available from: https://bmjpublichealth.bmj.com/content/3/1/e001836

40. Semenzato L, Botton J, Drouin J, et al. Chronic diseases, health conditions and risk of COVID-19-related hospitalization and in-hospital mortality during the first wave of the epidemic in France: a cohort study of 66 million people. Lancet Reg Health - Eur. 2021 Sep 1;8:100158.

41. Naouri D. COVID-19: third leading cause of death in France in 2020, while other major causes of death decline. Études et Résultats. 2022 Dec;1250:1–7 [Internet]. [cited 2024 Sep 19]. Available from: https://drees.solidarites-sante.gouv.fr/publications-en-anglais/covid-19-third-leading-cause-death-france-2020-while-other-major-causes

42. COVID-19 Data Explorer [Internet]. Our World in Data. [cited 2024 Sep 19]. Available from: https://ourworldindata.org/explorers/covid

43. World Health Organization (WHO). COVID-19 deaths | WHO COVID-19 dashboard [Internet]. datadot. [cited 2025 Jun 10]. Available from: https://data.who.int/dashboards/covid19/cases

44. McAlister FA, Parikh H, Lee DS, et al. Health Care Implications of the COVID-19 Pandemic for the Cardiovascular Practitioner. Can J Cardiol. 2023 Jun 1;39(6):716–25.

45. Gabet A, Grave C, Tuppin P, et al. Nationwide Initiation of Cardiovascular Risk Treatments During the COVID-19 Pandemic in France: Women on a Slippery Slope? Front Cardiovasc Med [Internet]. 2022 [cited 2023 Aug 2];9. Available from: https://www.frontiersin.org/articles/10.3389/fcvm.2022.856689

46. Wu J, Mamas MA, Mohamed MO, et al. Place and causes of acute cardiovascular mortality during the COVID-19 pandemic. Heart. 2021 Jan 1;107(2):113–9.

47. Shoaib A, Van Spall HGC, Wu J, et al. Substantial decline in hospital admissions for heart failure accompanied by increased community mortality during COVID-19 pandemic. Eur Heart J Qual Care Clin Outcomes. 2021 Jul 21;7(4):378–87.

48. Betai D, Ahmed AS, Saxena P, et al. Gender Disparities in Cardiovascular Disease and Their Management: A Review. Cureus [Internet]. 2024 May 5 [cited 2024 Sep 19];16(5). Available from: https://www.cureus.com/articles/245628-gender-disparities-in-cardiovascular-disease-and-their-management-a-review

49. López Ferreruela I, Obón Azuara B, Malo Fumanal S, et al. Gender inequalities in secondary prevention of cardiovascular disease: a scoping review. Int J Equity Health. 2024 Jul 23;23(1):146.

50. Grave C, Gabet A, Cinaud A, et al. Nationwide time trends in patients hospitalized for acute coronary syndrome: a worrying generational and social effect among women. Eur J Prev Cardiol. 2024 Jan 5;31(1):116–27.

51. Fouillet A, Cadillac M, Riviera C, et al. Leading causes of death in France in 2022 and recent trends [Internet]. Bull Épidémiol Hebd. 2024;(18):388–411. [cited 2024 Oct 10]. Available from: http://beh.santepubliquefrance.fr/beh/2024/18/2024_18_1.html

52. Boisguérin B, Gaimar L. En 2022, la baisse du nombre de lits en état d’accueillir des patients s’accentue | Direction de la recherche, des études, de l’évaluation et des statistiques [Internet]. [cited 2025 Jan 6]. Available from: https://drees.solidarites-sante.gouv.fr/publications-communique-de-presse/etudes-et-resultats/en-2022-la-baisse-du-nombre-de-lits-en-etat

53. Bates BA, Akhabue E, Nahass MM, et al. Validity of International Classification of Diseases (ICD)-10 Diagnosis Codes for Identification of Acute Heart Failure Hospitalization and Heart Failure with Reduced Versus Preserved Ejection Fraction in a National Medicare Sample. Circ Cardiovasc Qual Outcomes. 2023 Feb;16(2):e009078.

54. Frolova N, Bakal JA, McAlister FA, et al. Assessing the use of international classification of diseases-10th revision codes from the emergency department for the identification of acute heart failure. JACC Heart Fail. 2015 May;3(5):386–91.

