## Supplemental for "Hospitalisation for acute heart failure and in-hospital mortality before, during, and after the COVID-19 pandemic in France: A Nationwide cohort study from 2013 to 2024"

### Supplemental Text 1: Mapping algorithm used to define acute heart failure hospitalisation.

The algorithms of The Healthcare Expenditures and Conditions Mapping G11 version[1] were used to define Acute Heart Failure (AHF) hospitalisation. These algorithms have been subject to external expertise[1] and have already been used in a previous study.[2] The ICD-10 codes used and the data sources are detailed below:

***"Acute Heart Failure (MCV_ICA_AIG)***
*Patients hospitalised in care facilities (MCO) in year n for heart failure (as the principal diagnosis [PD] in one of the Medical Units [RUMs]) or for acute complications of heart failure (hypertensive heart disease with heart failure, hypertensive heart and renal disease with or without specification, congestive liver, or acute pulmonary oedema) (as the PD in one of the RUMs), with either heart failure as an associated diagnosis (AD) or related diagnosis (RD).*

***ICD-10 codes used:***

- ***PMSI (heart failure):*** *I50 (Heart failure)*
- ***PMSI (complications):***
  - *I11.0 (Hypertensive heart disease with congestive heart failure)*
  - *I13.0 (Hypertensive heart and renal disease with heart failure)*
  - *I13.2 (Hypertensive heart and renal disease with both (congestive) heart failure and renal failure)*
  - *I13.9 (Hypertensive heart and renal disease, unspecified)*
  - *K76.1 (Chronic passive congestion of the liver)*
  - *J81 (Pulmonary oedema)*

### Supplemental Analysis 1: COVID-19 mortality among patients who died during AHF hospitalisation.

To quantify the proportion of in-hospital deaths during AHF hospitalisation directly attributable to COVID-19, the causes of death were analysed. COVID-19-attributed deaths were defined using the ICD-10 codes U07.1 and U07.2 as the leading causes of death.[3] This analysis was conducted only for the years 2020 to 2022, as causes of death were not available for the years 2023 and 2024.

Between 2020 and 2022, a total of 2,738 AHF-hospitalised patients died during the hospitalisation, with COVID-19 identified as the leading cause of death, of whom 1,309 (47.8%) were female.

In 2020, 929 COVID-19-related deaths were recorded, exceeding the 828 estimated excess in-hospital deaths for that year. Conversely, in 2021 and 2022, the numbers of COVID-19-related deaths – 957 and 852, respectively – were lower than the estimated excess in-hospital deaths, which were 1,625 for 2021 and 2,427 for 2022.

In 2021 and 2022, COVID-19 mortality was lower among females than among males, while the estimated excess in-hospital mortality during these years was higher among females than among males (see Table 1, Supplemental Table 1). While COVID-19-related mortality exceeded the estimated excess in-hospital mortality in 2020, it declined over time, accounting for only 35.1% of the estimated excess mortality in 2022 (Supplemental Table 1).

**Supplemental Table 1: COVID-19 mortality among patients hospitalised for acute heart failure and relative proportion compared to estimated excess mortality, France, 2020–2022.**

| COVID-19 deaths among AHF hospitalised patients* | Year | | |
|---|---|---|---|
| | **2020** | **2021** | **2022** |
| **Total** | **929 (112,3%)** | **957 (58,9%)** | **852 (35.1%)** |
| Females | 455 (113,2%) | 455 (48,8%) | 399 (27.1%) |
| Males | 474 (111.5%) | 502 (72.6%) | 453 (47.5%) |

*Number of patients who died with COVID-19 as the leading cause of death during hospitalisation for acute heart failure (relative proportion compared to excess in-hospital deaths estimated).
Each cell contains the COVID-19 mortality among AHF hospitalised patients, with the relative proportion compared to excess deaths estimated. For instance, in 2022, 399 females hospitalised for acute heart failure died with COVID-19 as the leading cause of death; this amount represents 27.1% of the 1473 excess deaths estimated for females this year (see Table 1).

### Supplemental Figure 1: Expected and observed AHF hospitalisations by year, France, 2013–2024.

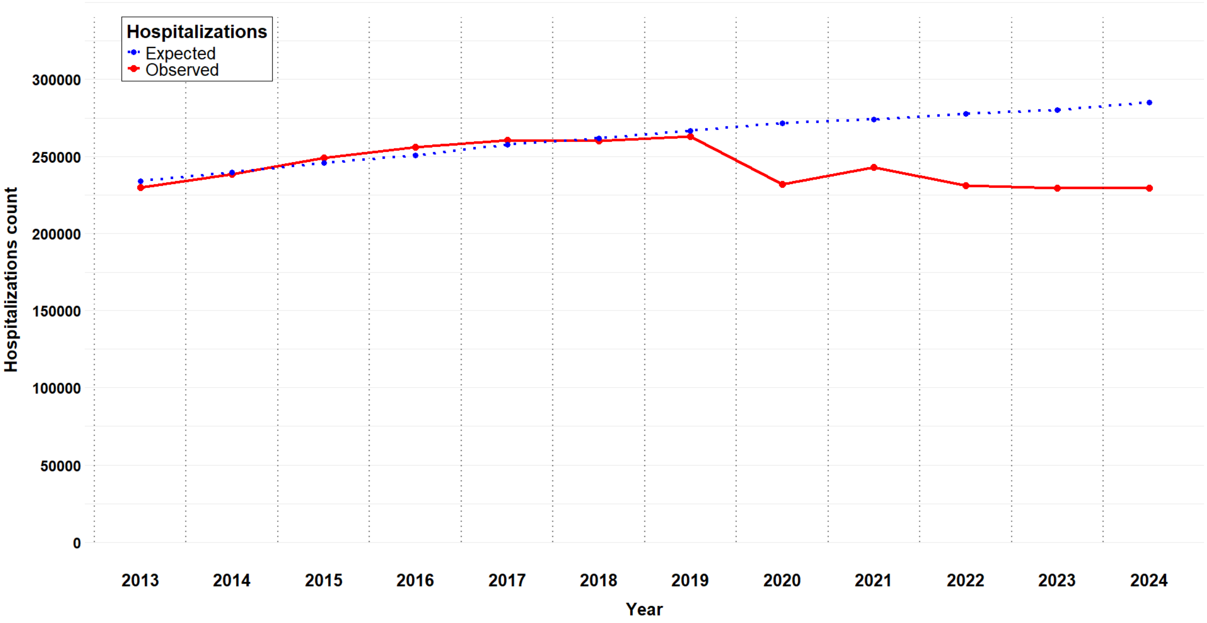


For each year from 2013 to 2024, the red curve represents the observed AHF hospitalisations in France. The blue curve represents the AHF hospitalisations predicted by the Poisson regression model described in the main article, with 2013–2019 as the reference period.

### Supplemental Table 2: Overview of the linear predictors used in the Poisson regression model to estimate hospitalisation count, with corresponding AIC and rationale, reference period 2013–2019.

| Linear predictor | AIC | Rationale |
|---|---|---|
| Year + Sex + $Age_c$ | **1,021,358** | Year, sex, and age were included as continuous variables; estimates average linear effects. |
| Year + Sex + $Age_c$ + log(Population) | **85,578** | Population was added as an offset to account for changes in population structure over time. |
| Year + Sex + $Age_f$ + log(Population) | **35,986** | Age was modelled as a categorical variable to capture non-linear age effects. |
| Year + Sex · $Age_f$ + log(Population) | **13,243** | Interaction between sex and age was included to account for varying effects of sex across age groups. |

$Age_c$ – age coded as a continuous variable; $Age_f$ – age coded as a factor (categorical variable)

### Supplemental Analysis 2: Impact of the intra-annual periodicity on estimation of AHF hospitalisation count.

To explore the seasonal periodicity of hospitalisations and assess its impact on the estimation of the decline in hospitalisations during the pandemic period, we investigated two additional models. The first model, henceforth referred to as the "Sinus model", utilized monthly granularity (month) and incorporated harmonic terms, $sin(2\pi \cdot Month_l/12)$ and $cos(2\pi \cdot Month_l/12)$, to account for annual periodicity, using the same notations as in the main text and $Month_l$ ($l \in [\![1, \ldots, 12]\!]$) the month number within a year. The analytical expression of this model is as follows:

$$\log(\widehat{eH}_{i,j,k,l}) = \log\left(\text{Population}_{i,j,k}\right) + \beta_{h_0} + \beta_{h_1} \cdot \text{Sex}_i + \beta_{h_2} \cdot \text{Age}_j + \beta_{h_3} \cdot \text{Year}_k + \beta_{h_4} \cdot \text{Sex}_i \cdot \text{Age}_j + \beta_{h_5} \cdot sin\left(\frac{2\pi \cdot Month_l}{12}\right) + \beta_{h_6} \cdot cos\left(\frac{2\pi \cdot Month_l}{12}\right)$$

As shown in Supplemental Figure 2, this periodic model effectively captures the seasonal trend in hospitalisations during the pre-pandemic period. Moreover, it highlights the sharp decline in hospitalisations starting with the onset of the pandemic in 2020, which persisted through 2024.

**Supplemental Figure 2. Expected and observed hospitalisations by month, Sinus model, France, 2013–2024.**

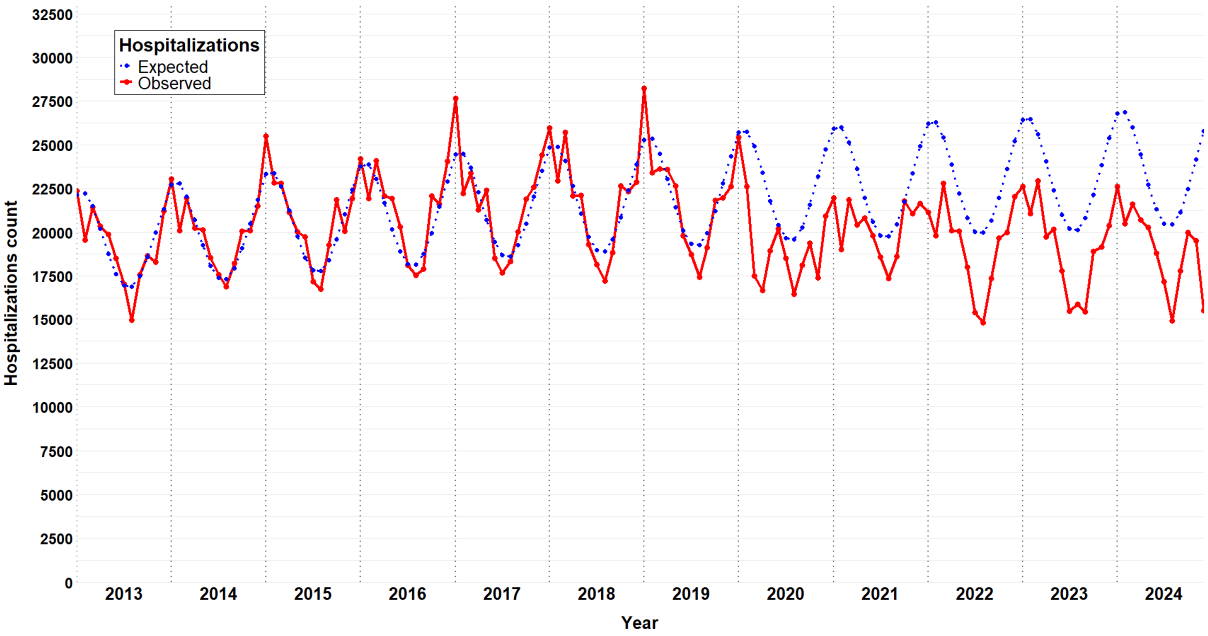


For each year from 2013 to 2024, the red curve represents the monthly observed AHF hospitalisations in France. The blue curve represents the AHF hospitalisations predicted by the "Sinus" regression model described before, with 2013–2019 as the reference period.

However, this model does not perfectly capture extreme variations, such as the decline in hospitalisations during the summer or the peak at the beginning of the year. Therefore, a second model with finer temporal granularity (week) and greater flexibility (spline with 30 degrees of freedom) was also evaluated. This second model is henceforth referred to as the "Spline model." The analytical expression of this model is as follows:

$$\log(\widehat{eH}_{i,j,k,m}) = \log\left(\text{Population}_{i,j,k}\right) + \beta_{h_0} + \beta_{h_1} \cdot \text{Sex}_i + \beta_{h_2} \cdot \text{Age}_j + \beta_{h_3} \cdot \text{Year}_k + \beta_{h_4} \cdot \text{Sex}_i \cdot \text{Age}_j + \beta_{h_5} \cdot \text{ns}(week_m\text{ , df=30})$$

As shown in Supplemental Figure 3, this new model appears to be more accurate in capturing the extremes of seasonal variations. It also highlights a significant decline in hospitalisations during the pandemic period, which was particularly abrupt in the spring of 2020, during the first pandemic wave, and shows that these disruptions persisted until 2024.

**Supplemental Figure 3. Expected and observed hospitalisations by week, Spline model, France, 2013–2024.**

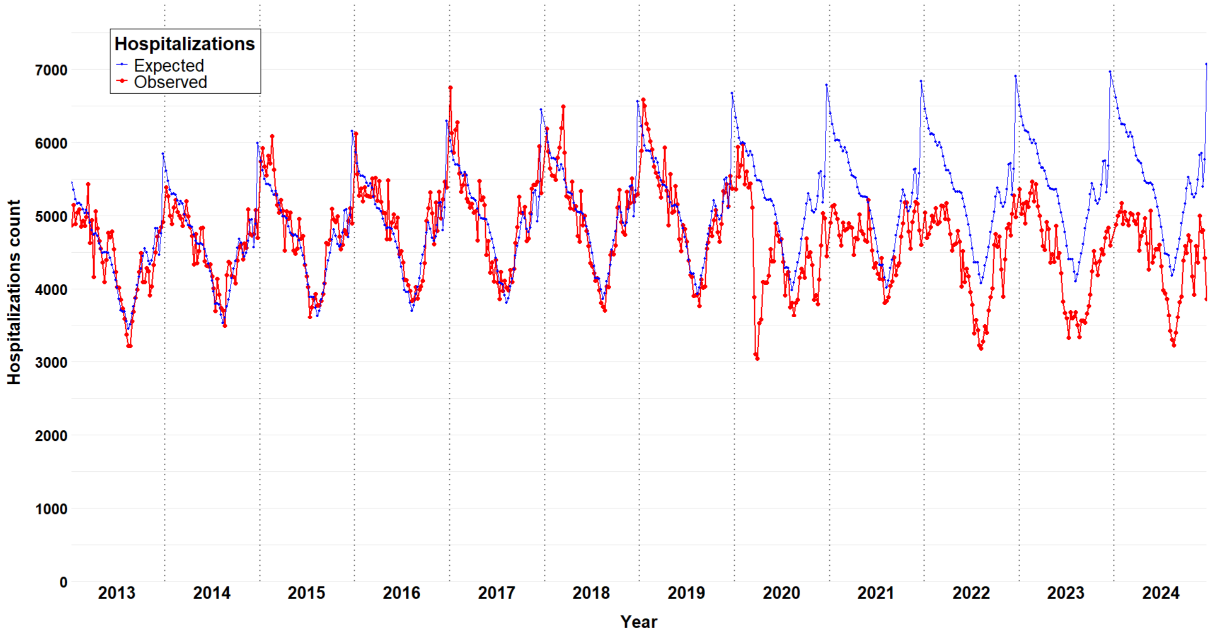


For each year from 2013 to 2024, the red curve represents the number of AHF hospitalisations observed weekly in France. The blue curve represents the corresponding numbers of AHF hospitalisations predicted by the "Spline" regression model described before, using years 2013–2019 as the reference period.

As shown in Supplemental Figure 4, the estimates of the reduction in hospitalisations during the pandemic period using both periodic models are close to those obtained with the initial Poisson model presented in the main article. However, the periodic models require additional methodological decisions regarding temporal granularity, the number of harmonics, or degrees of freedom, without significantly impacting the final results. In the interest of parsimony, we therefore chose to retain the simpler Poisson model for the primary analysis.

**Supplemental Figure 4. Difference between observed and expected hospitalisations by model and by year, France, 2020–2024.**

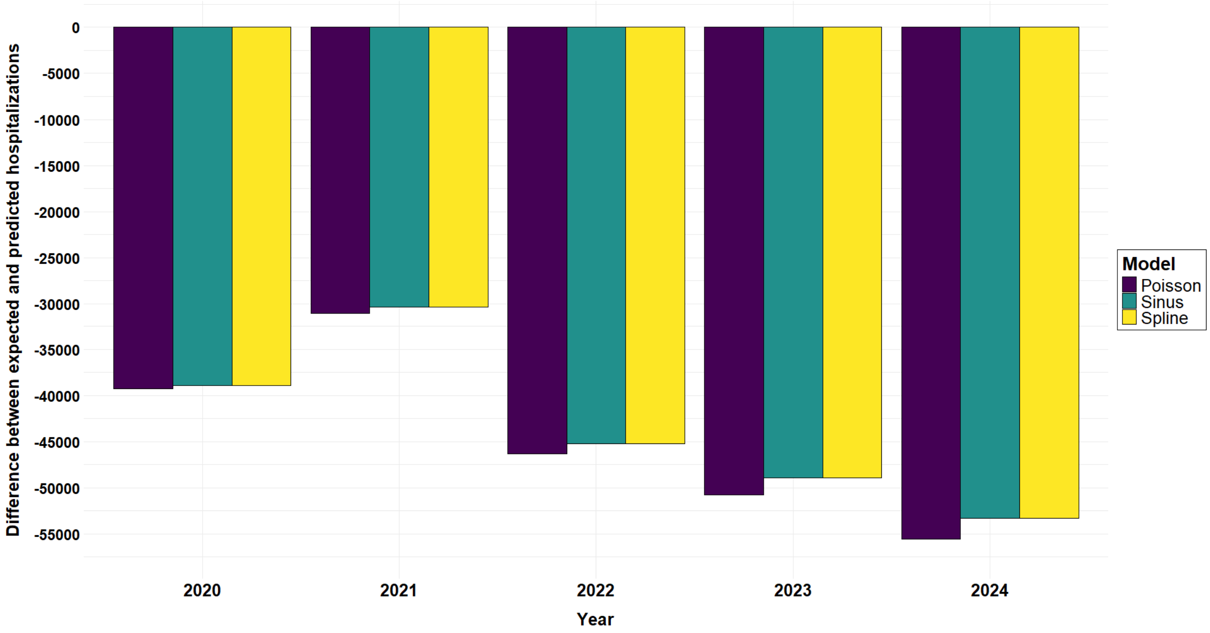


For each model and each year, the bar represents the difference between the observed number of AHF hospitalisations and those predicted by the corresponding model. For example, in 2020, the drop in hospitalisations estimated with the Poisson regression model presented in the main article is 39,268, representing a 14.5% decrease compared to the expected level. This is close to the 38,937-drop estimated with the 'Spline' model, which represented a 14.4% decrease compared to the expected number.

**Supplemental Figure 5. Population distribution (sex and age) of AHF hospitalisations and estimated drop in hospitalisations, France, 2020–2024.**

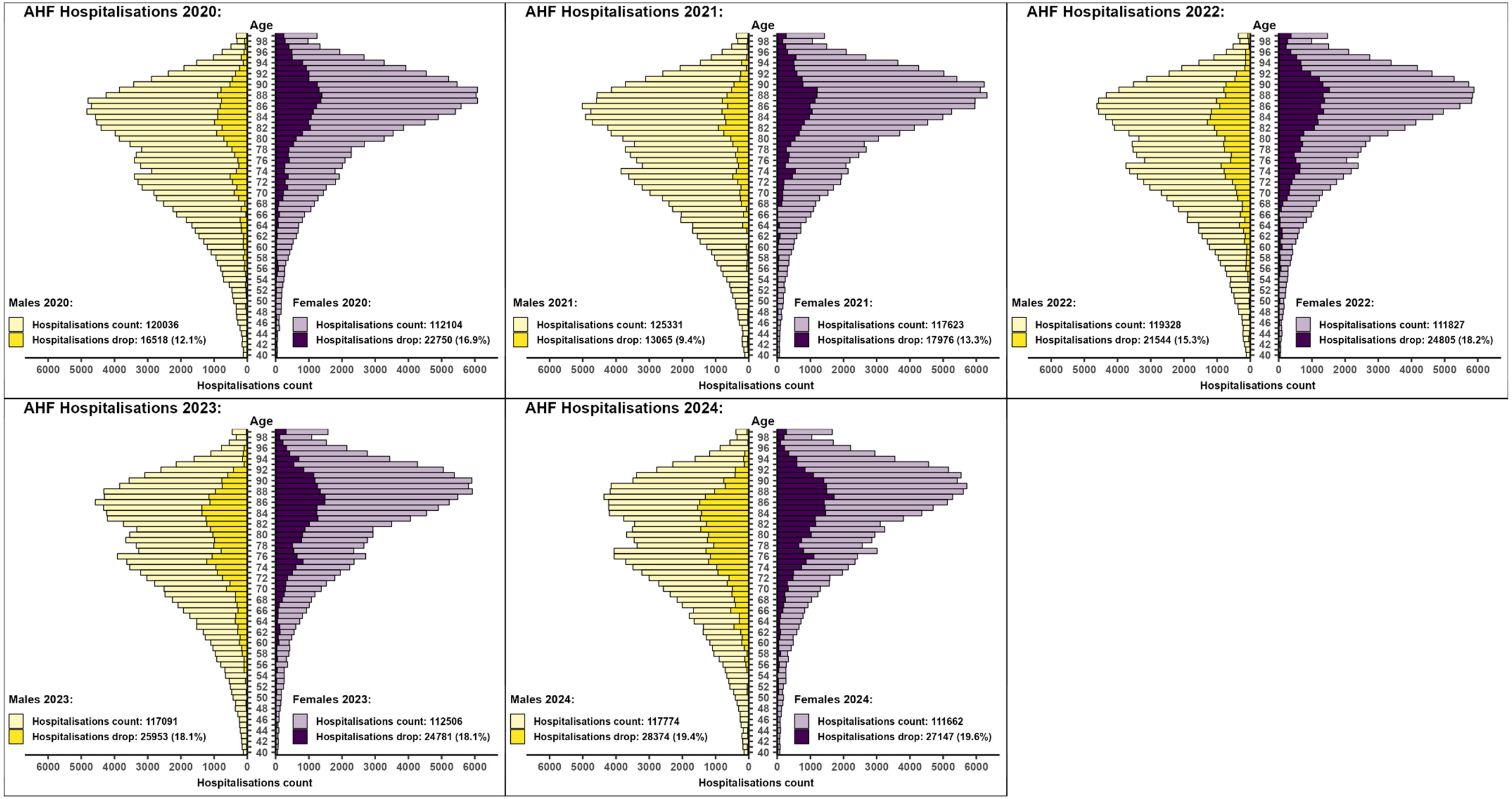


For each year between 2020 and 2024, age pyramids represent the observed values for AHF hospitalisations in light bars, and the estimated drop in hospitalisations in dark bars. The drop in hospitalisations corresponds to the number of hospitalisations missing to reach the expected level based on the pre-pandemic trend. For example, in 2020 (first pyramid), for males (yellow), around 5,000 patients aged 85 years were hospitalised for AHF (light yellow), about 1,000 fewer than expected (dark yellow).

**Supplemental Figure 6. Population distribution (sex and age) of in-hospital deaths during AHF hospitalisations and estimated excess mortality, France, 2020–2024.**

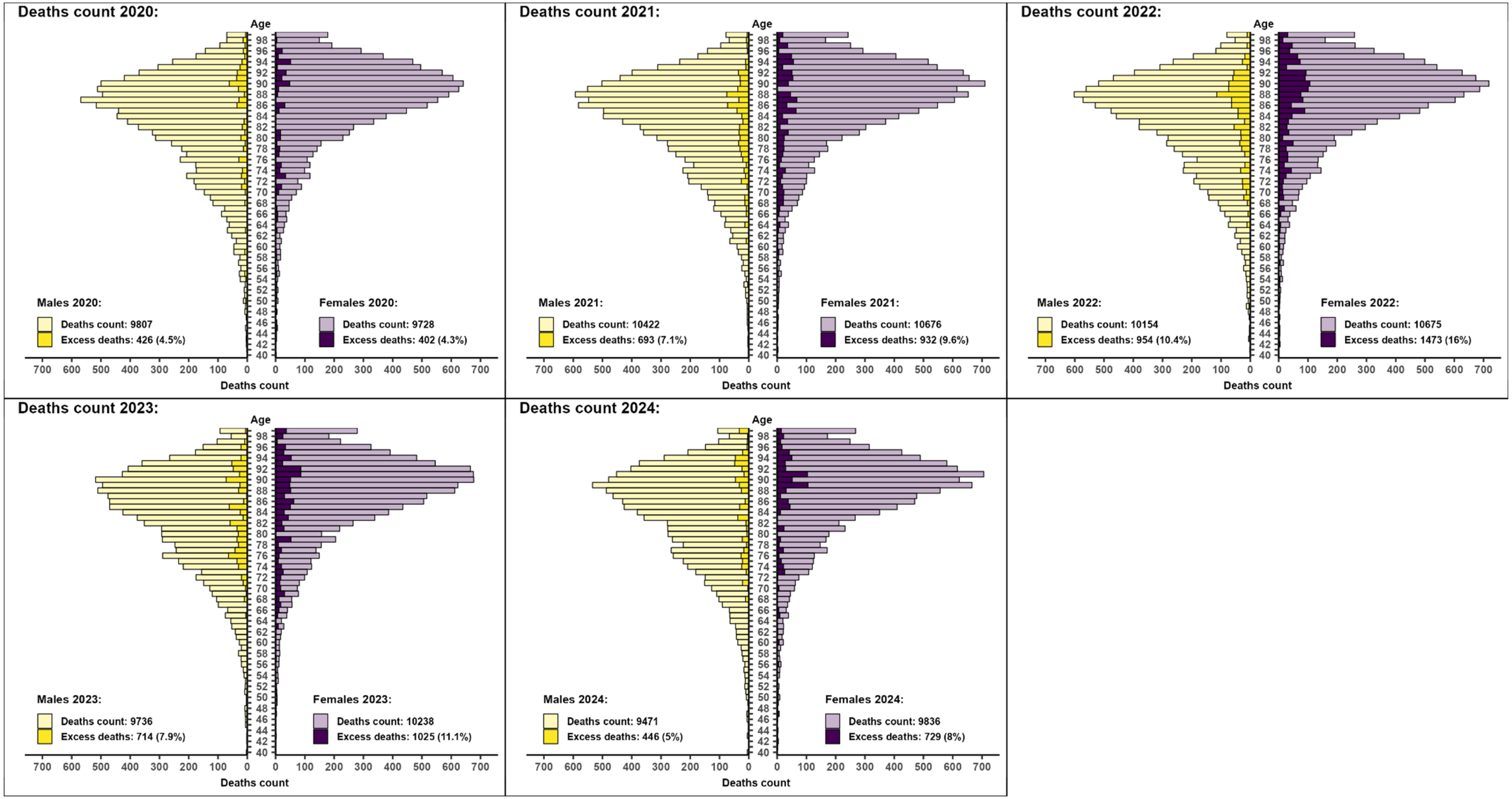


For each year between 2020 and 2024, an age pyramid represents the observed values for deaths during AHF hospitalisations in light bars, and the estimated excess deaths in dark bars. Excess deaths refer to the number of deaths above the expected level based on pre-pandemic trends. For example, in 2022, 700 females aged 90 (light purple) years died during AHF hospitalisation, about 100 more than expected (dark purple).

**Supplemental Analysis 3: Changes in all-cause hospitalisations during the pandemic period.**

To assess whether the decline in hospitalisations was specific to AHF, a supplemental analysis was conducted on all-cause hospitalisations (excluding AHF). All hospitalisations with at least one overnight stay between 2013 and 2024 were considered (n=111,720,823). As shown in Supplemental Figure 7, a substantial decrease has been observed since the onset of the COVID-19 pandemic in 2020. No catch-up phenomenon was observed in the following years, and annual hospitalisation levels never returned to pre-pandemic levels.
The same modelling approach described in the main article was used to estimate the magnitude of this disruption. A total reduction of 4,159,186 [4,121,232; 4,195,424] hospitalisations was estimated between 2020 and 2024, corresponding to 8.71% [8.64%; 9.78%] fewer hospitalisations than expected based on pre-pandemic trends. The decline peaked in 2020, with a reduction of 1,118,422 [1,110,453; 1,125,841] hospitalisations, and decreased to 710,227 [699,070; 720,828] in 2024 (Supplemental Table 3).

**Comparison of the drop in hospitalisations, all causes vs AHF.**

To compare the magnitude of the decline in AHF hospitalisations relative to all-cause hospitalisations, the ratio of the reduction was calculated. Over the entire pandemic period (2020−2024), AHF hospitalisations decreased by 16.1%, compared to an 8.7% reduction for all-cause hospitalisations, yielding a ratio of 1.9. Specifically, the ratio was 1.3 in 2020 and increased to 2.6 in 2024. Notably, while the decline in hospitalisations diminished over time in the general population, it worsened among AHF patients.

**Conclusion.**

The impact of the pandemic period on hospitalisations was not limited to AHF but also affected the general population, with a brutal and lasting decrease in hospitalisations. However, when examining the magnitude and dynamics of these disruptions, AHF hospitalisations were disproportionately affected. This gap widened over time, ultimately leading to an overall impact on AHF hospitalisations that was nearly twice as severe as that observed in the general population.

**Supplemental Figure 7. Annual standardised count of hospitalisations in million, France, 2020–2024**

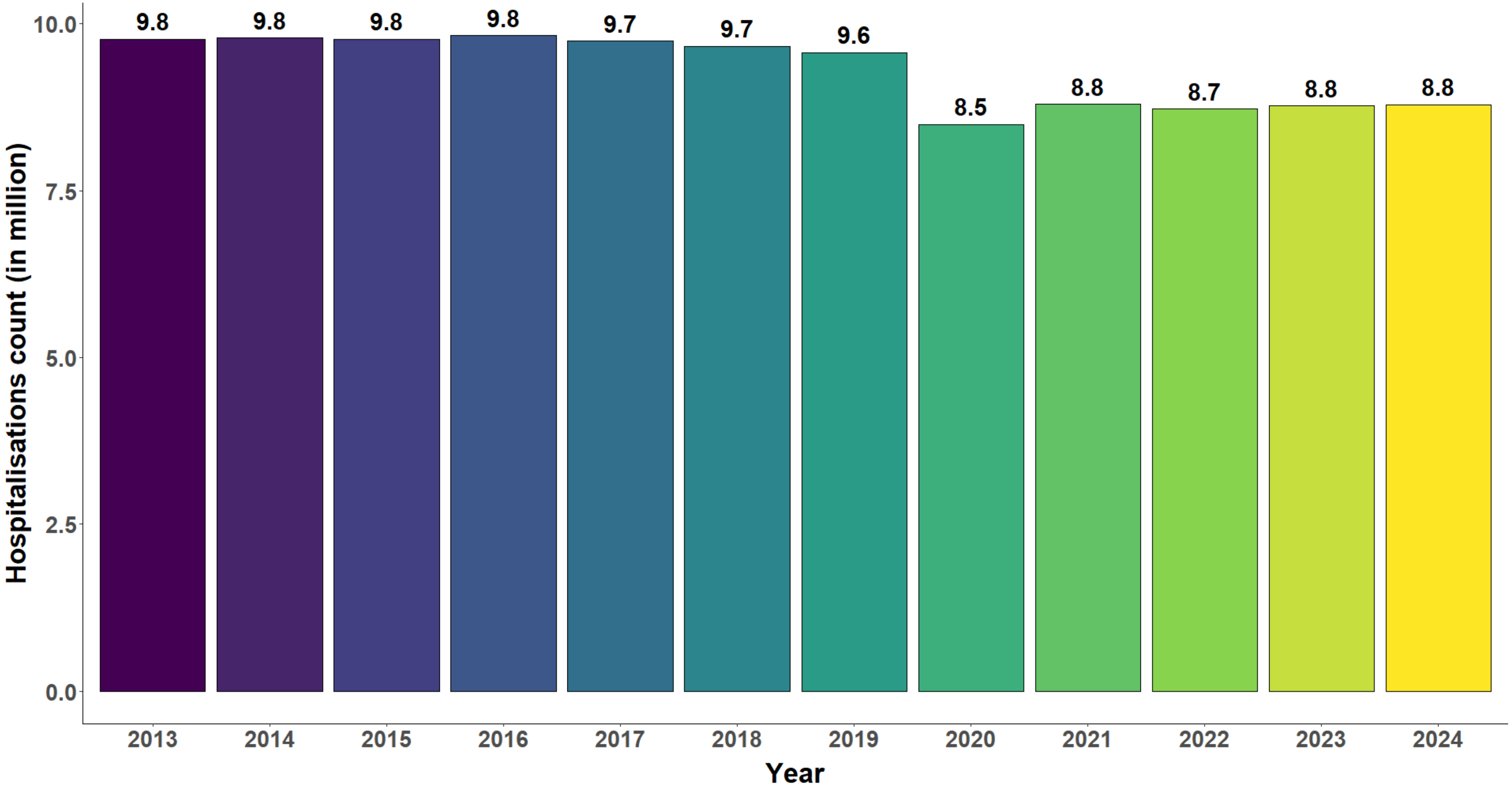


For each year between 2013 and 2024, the bar represents the cumulative number of standardised hospitalisations. In order to standardise analyses, the average population by age and sex between 2013 and 2024 was the reference.

### Supplemental Table 3: Differences (observed - expected) in the numbers of all cause hospitalisations, France, 2020−2024

| Difference (observed – expected) * | Year | | | | |
|---|---|---|---|---|---|
| | 2020 | 2021 | 2022 | 2023 | 2024 |
| **Number of hospitalisations** | -1,118,422 [-1,125,841; -1,110,453]<br>-11.64% [-11.71%; -11.56%] | -774,966 [-782,851; -766,724]<br>-8.1% [-8.17%; -8.01%] | -818,930 [-828,014; -809,342]<br>-8.58% [-8.67%; -8.49%] | -736,642 [-746,875; -726,655]<br>-7.74% [-7.85%; -7.65%] | -710,227 [-720,828; -699,070]<br>-7.48% [-7.58%; -7.37%] |
| **Females** | -605,892 [-611,358; -600,738]<br>-11.89% [-11.99%; -11.79%] | -387,775 [-393,719; -381,808]<br>-7.67% [-7.78%; -7.56%] | -399,856 [-406,047; -393,353]<br>-7.96% [-8.08%; -7.84%] | -358,714 [-365,939; -351,719]<br>-7.2% [-7.34%; -7.06%] | -327,391 [-334,845; -319,639]<br>-6.61% [-6.75%; -6.45%] |
| **Males** | -512,529 [-517,862; -506,883]<br>-11.35% [-11.46%; -11.23%] | -387,190 [-393,092; -381,473]<br>-8.58% [-8.7%; -8.46%] | -419,074 [-425,331; -412426]<br>-9.27% [-9.4%; -9.13%] | -377,928 [-385,126; -370,515]<br>-8.35% [-8.5%; -8.19%] | -382,836 [-390,430; -374,909]<br>-8.42% [-8.58%; -8.26%] |

*Differences expressed as: mean difference [95% confidence interval] (disruption of the evolution relative to the expected trend, i.e., difference / expected number × 100); reference period for estimating expected numbers = years 2013−2019.

The negative difference between observed and expected AHF hospitalisations corresponds to a drop in hospitalisations.

### References


1. Méthodehttps://www.assurance-maladie.ameli.fr/etudes-et-donnees/par-theme/pathologies/cartographie-assurance-maladie/methode-cartographie-pathologies-depenses-assurance-maladie (20 September 2024)

2. Rachas A, Gastaldi-Ménager C, Denis P, Barthélémy P, Constantinou P, Drouin J, Lastier D, Lesuffleur T, Mette C, Nicolas M, Pestel L, Rivière S, Tajahmady A, Gissot C, Fagot-Campagna A. The Economic Burden of Disease in France From the National Health Insurance Perspective. *Med Care* 2022;**60**:655–664.

3. Fouillet A, Cadillac M, Riviera C, Coudin E. Leading causes of death in France in 2022 and recent trends. *Bull Épidémiol Hebd. 2024;(18):388-411.* http://beh.santepubliquefrance.fr/beh/2024/18/2024_18_1.html (10 October 2024)
